# Tuberculosis healthcare service disruptions during the COVID-19 pandemic in Brazil, India and South Africa: A model-based analysis of country-level data

**DOI:** 10.1101/2024.05.16.24307503

**Authors:** Abigail K. de Villiers, Muhammad Osman, Claudio J. Struchiner, Anete Trajman, Dheeraj Tumu, Vaibhav V. Shah, Guilherme L. Werneck, Layana C. Alves, Megha Choudhary, Sunita Verma, Sanjay K. Mattoo, Sue-Ann Meehan, Urvashi B. Singh, Anneke C. Hesseling, Florian M. Marx

**Affiliations:** Desmond Tutu TB Centre, Department of Paediatrics and Child Health, Faculty of Medicine and Health Sciences, Stellenbosch University, South Africa; South African Centre for Epidemiological Modelling and Analysis, Stellenbosch University, Stellenbosch, South Africa; Department of Infectious Diseases and Tropical Medicine, Heidelberg University Hospital, Heidelberg, Germany; School of Human Sciences, Faculty of Education, Health and Human Sciences, University of Greenwich, United Kingdom; School of Applied Mathematics, Fundação Getúlio Vargas, Rio de Janeiro, Brazil; Department of Internal Medicine, Medical School, Federal University of Rio de Janeiro, Brazil; Central TB Division, National TB Elimination Program, Ministry of Health, Government of India; Department of Epidemiology, State University of Rio de Janeiro, Brazil; Collective Health Institute, Federal University of Bahia, Salvador, Brazil; Department of Microbiology, All India Institute of Medical Sciences, New Delhi, India

## Abstract

Tuberculosis (TB) is the leading infectious disease cause of death worldwide. In recent years, stringent measures to contain the spread of SARS-CoV-2 have led to considerable disruptions of healthcare services for TB in many countries. The extent to which these measures have affected TB testing, treatment initiation and outcomes has not been comprehensively assessed. We aimed to estimate TB healthcare service disruptions occurring during the COVID-19 pandemic in Brazil, India, and South Africa. We obtained country-level TB programme and laboratory data and used autoregressive integrated moving average (ARIMA) time-series models to estimate healthcare service disruptions with respect to TB testing, treatment initiation, and treatment outcomes. We quantified disruptions as the percentage difference between TB indicator data observed during the COVID-19 pandemic compared with values for a hypothetical no-COVID scenario, predicted through forecasting of trends during a three-year pre-pandemic period. Annual estimates for 2020-2022 were derived from aggregated monthly data. We estimated that in 2020, the number of bacteriological tests conducted for TB diagnosis was 24.3% (95% uncertainty interval: 8.4%;36.6%) lower in Brazil, 27.8% (19.8;3 4.8%) lower in India, and 32.0% (28.9%;34.9%) lower in South Africa compared with values predicted for the no-COVID scenario. TB treatment initiations were 17.4% (13.9%;20.6%) lower than predicted in Brazil, 43.3% (39.8%;46.4%) in India, and 27.0% (15.2%;36.3%) in South Africa. Reductions in 2021 were less severe compared with 2020. The percentage deaths during TB treatment were 13.7% (8.1%; 19.7%) higher than predicted in Brazil, 1.7% (−8.9%;14.0%) in India and 21.8% (7.4%;39.2%) in South Africa. Our analysis suggests considerable disruptions of TB healthcare services occurred during the early phase of the COVID-19 pandemic in Brazil, India, and South Africa, with at least partial recovery in the following years. Sustained efforts to mitigate the detrimental impact of COVID-19 on TB healthcare services are needed.

## Introduction

Tuberculosis (TB) continues to be a serious threat to population health globally. In 2022, an estimated 10.6 million individuals worldwide developed TB and 1.6 million TB-related deaths occurred [1] making TB the single most common infectious disease cause of death worldwide [2]. The SARS-CoV-2 pandemic exacerbated TB as a public health concern [3]. The World Health Organization reported that in 2020, 1.4 million fewer people received TB treatment, compared with those reported in 2019 [4, 5]. Low- and middle-income countries (LMICs), which account for 95% of all TB deaths worldwide, were severely affected by the social and economic consequences of the COVID-19 pandemic and related response measures [6].

During lockdowns implemented to contain SARS-CoV-2 transmission, individuals faced difficulties in accessing TB healthcare services due to movement restrictions, increased financial constraints and elevated fear of contracting COVID-19 in healthcare settings [7, 8]. Healthcare service delivery was negatively impacted as human and financial resources were redirected towards addressing the demands of the COVID-19 pandemic. Staff shortages due to COVID-19 illness further impacted TB testing and treatment [9, 10]. Consequently, these disruptions may have driven delayed and missed TB diagnoses, reduced numbers of people initiating TB treatment, and increased rates of unfavourable TB treatment outcomes [11].

Several countries have emphasized disruptions to TB healthcare services due to efforts to reduce the spread of SARS-CoV-2. A survey of Global Fund-supported programmes across 106 countries found that 78% of TB programmes experienced substantial disruptions, with 17% experiencing high or very high levels of disruption [12]. According to WHO data from March 2021, COVID-19 disruptions had a significant impact on TB care in over 84 countries; it was estimated that these disruptions could be as high as 25%-50% [13].

A review of studies on COVID-19 impact on TB published in 2021 found that substantial disruptions to TB services and increased vulnerability to TB had occurred [14]. Early evaluations quantifying country-level disruptions to TB healthcare services in Brazil [15, 16], India [17] and South Africa [18, 19] have been conducted [12]. However, these studies were limited in capturing the full extent of TB healthcare service disruptions as they relied on relatively short observation periods conducted before and after comparisons of indicators without taking seasonal or annual trends into account, or did not include an assessment of data on TB treatment outcomes.

This study formed part of the IMPAC_19_T_B_ project, a large multi-national research project on the epidemiological impact and intersection of the COVID-19 and tuberculosis pandemics in Brazil, Russia, India and South Africa [20]. Using country-level TB programmatic and laboratory data, we aimed to estimate the extent of TB healthcare service disruptions from the onset of the COVID-19 pandemic in Brazil, India, and South Africa.

## Methods

### Study setting

This study focused on Brazil, India and South Africa, three countries which rank among the 30 countries with the highest TB burden in terms of estimated incident TB cases and were also severely affected by the COVID-19 pandemic [1]. Key indicator data for COVID-19 and TB for the three countries are provided in Table 1.

**Table 1.** Population size, TB incidence and COVID-19 cases and deaths per country.

|  | **Population size (2022)**<br>[21] | **Total TB incidence (2022)**<br>[1] | **HIV-positive TB incidence (2022)**<br>[1] | **COVID-19 cases – cumulative\***<br>[22] | **COVID-19 deaths\***<br>[22] |
| --- | --- | --- | --- | --- | --- |
| **Brazil** | 215,313,498 | 105,000<br>(89,000-121,000) | 19,000<br>(16,000-23,000) | 38,210,864 | 708,638 |
| **India** | 1,417,173,173 | 2,820,000<br>(2,390,000-3,280,000) | 48,000<br>(40,000-55,000) | 45,020,333 | 533,049 |
| **South Africa** | 59,893,885 | 280,000<br>(182,000-398,000) | 152,000<br>(99,000-217,000) | 4,076,463 | 102,595 |

\*29<sup>th</sup> January 2020- 12th January 2024.

### Indicators and data sources

We assessed country-level trends in selected key TB indicators as proxy information to estimate the extent of TB healthcare service disruptions during the COVID-19 pandemic (Table 2).

**Table 2.** TB indicators evaluated.

| TB indicator | Description |
| --- | --- |
| **TB testing data** |  |
| Number of TB tests conducted | Number of bacteriological TB tests* conducted |
| Test positivity | Number of bacteriological TB tests with a positive test result divided by the number of bacteriological tests conducted |
| **TB treatment initiation data** |  |
| Number of people initiating TB treatment | Number of individuals reported to have initiated TB treatment; based on case notification data recorded in TB treatment registers |
| **TB treatment outcome data<sup>†</sup>** |  |
| Percentage unfavourable treatment outcomes | Number of patients who had a standard treatment outcome other than treatment success (i.e. either failure, death, or loss to follow-up) divided by the total number of patients with reported TB treatment outcomes |
| Percentage treatment failure | Number of patients with the standard treatment outcome 'treatment failure' divided by the total number of patients with reported TB treatment outcomes |
| Percentage death during treatment | Number of patients with the standard treatment outcome 'death' divided by the total number of patients with reported TB treatment outcomes |
| Percentage loss to follow-up | Number of patients with the standard treatment outcome 'loss to follow-up from treatment' divided by the total number of patients with reported TB treatment outcomes |

\* TB test data represent numbers of bacteriological tests conducted per unit time. For Brazil and South Africa bacteriological tests included WHO-recommended rapid diagnostics (WRDs) as per National TB Programme guidelines whilst for India, tests included sputum smear microscopy and WRDs.
<sup>†</sup> Percentage treatment outcomes refer to individuals who initiated TB treatment in that month (outcome cohort) excluding outcomes reported as unknown or not evaluated.

We hypothesized that numbers of bacteriological tests conducted for the diagnosis of TB may be impacted by reduced access to TB testing and/or reduced testing capacity during the COVID-19 pandemic. Numbers of people initiating TB treatment may have been impacted by either reduced access to TB testing, or losses and delays prior to the uptake of treatment. Standard TB treatment outcomes may be impacted by diagnostic delays as well as reduced access, supervision, and support during treatment. Data for these key indicators were collected from in-country routine programmatic clinical and laboratory data sources (Table 3).

**Table 3.** TB programmatic available time periods and data sources.

| **Data type** | **Country** | **Time period** | **Source** | **Date accessed** |
| --- | --- | --- | --- | --- |
| **TB testing** | Brazil | 2017 - 2022 | Tuberculosis Rapid Test Network (RTR-TB, Rede de Testes Rápidos da Tuberculose) | 18/12/2022 |
|  | India | 2019 – 2022<sup>a</sup> | Nikshay Portal, Central TB division, Ministry of health and family welfare [23] | 02/08/2023 |
|  | South Africa | 2017 – 2022<sup>b</sup> | National Health Laboratory Service (NHLS) via National Institute for Communicable Diseases (NICD) | 04/07/2022 |
| **TB treatment initiation** | Brazil | 2017 - 2022 | Notifiable Diseases Information System, Ministry of Health [24] | 10/04/2023 |
|  | India | 2017 - 2022 | Nikshay Portal, Central TB division, Ministry of health and family welfare [23] | 02/08/2023 |
|  | South Africa | 2018 - 2021 | Health Informatics Directorate of the National Department of Health | 04/07/2022 |
| **TB treatment outcomes** | Brazil | 2017 - 2021 | Notifiable Diseases Information System, Ministry of Health [24] | 10/04/2023 |
|  | India | 2017 - 2021 | Nikshay Portal, Central TB division, Ministry of health and family welfare [23] | 02/08/2023 |
|  | South Africa | 2018 - 2020 | Health Informatics Directorate of the National Department of Health | 04/07/2022 |

<sup>a</sup> Quarterly-level estimates only
<sup>b</sup> until 06/2022

### Modeling approach

We used autoregressive integrated moving average (ARIMA) time-series models to investigate TB healthcare service disruptions with respect to TB testing, treatment initiation and outcomes during the first 2 years of the COVID-19 pandemic (April 2020-December 2021). ARIMA models are statistical models which are fitted to time-series data to predict (forecast) values for future points in time [25]. They combine auto-regression with moving averages to account for secular and seasonal growth or decline and random variation in the data. The ARIMA models were implemented in R studio (version 2022.02.2) using the *auto.arima()* function from the *forecast* package [26].

We estimated monthly healthcare service disruptions during the COVID-19 pandemic as the percentage difference between indicator data observed during the pandemic compared with values predicted through forecasting of pre-pandemic trends using reported data from a three-year (2017-2019) pre-COVID-19 period (no-COVID scenario).

While forecasting of pre-COVID trends referred to monthly (and quarterly) time points, we then derived aggregated annual estimates of percentage differences for each indicator as follows. We sampled 10,000 predicted values from the ARIMA model using the *simulate()* function

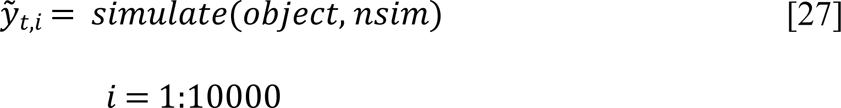

where 𝑦_𝑡,𝑖_represents the 𝑖^th^ sampled estimate of the predicted TB indicator value at time 𝑡 (calendar month), the *object* is the ARIMA model function and *nsim* the number of predicted time points *t*. We then calculated predicted annual estimates as the sums of monthly estimates

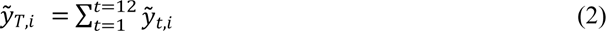

where 𝑡 represents the calendar month and 𝑇 the calendar year.

Estimates of annual percentage difference (observed vs. predicted) with 95% uncertainty intervals (UI) were then derived from the 10,000 sampled estimates using the following formulas

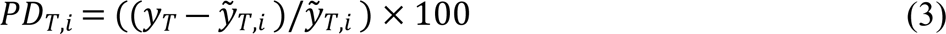

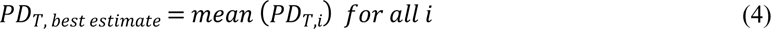

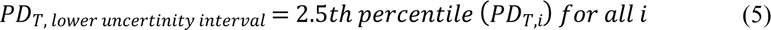

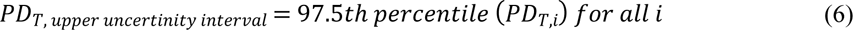

where 𝑃𝐷 represents the percentage difference, 𝑦 represents the observed value, 𝑦 represents the value predicted by the ARIMA model and 𝑇 represents the time (calendar year).

### Ethics

This was part of the IMPAC_19_T_B_ project approved by the Ethics Committee of the Institute of Social Medicine, University of the State of Rio de Janeiro, Brazil (4.784.355), the Ethics Committee of the All India Institute of Medical Sciences in India (IEC-305/07.04.2021, RP-21/2021) and the Health Research Ethics Committee of Stellenbosch University, South Africa (N21/05/013_COVID-19) and local/national TB program approvals were obtained. All analyses for this study were based on aggregated (de-identified) programmatic and laboratory data.

## Results

### Trends in TB testing, treatment initiation and outcomes during the pre-COVID-19 period (2017-2019)

Prior to the COVID-19 pandemic, between 2017 and 2019, the number of TB tests conducted had been slightly increasing over time in Brazil and India, while South Africa maintained stable test numbers with well-described seasonal short-term periodic decreases in December each year (Figure 1). The number of TB treatment initiations had been increasing in Brazil, India, and South Africa, with India showing the most substantial increase (Figure 1). The percentage of patients with unfavorable treatment outcomes had been increasing in Brazil, stable or slightly decreasing in India, and stable in South Africa (Figure 2).

**Figure 1.**
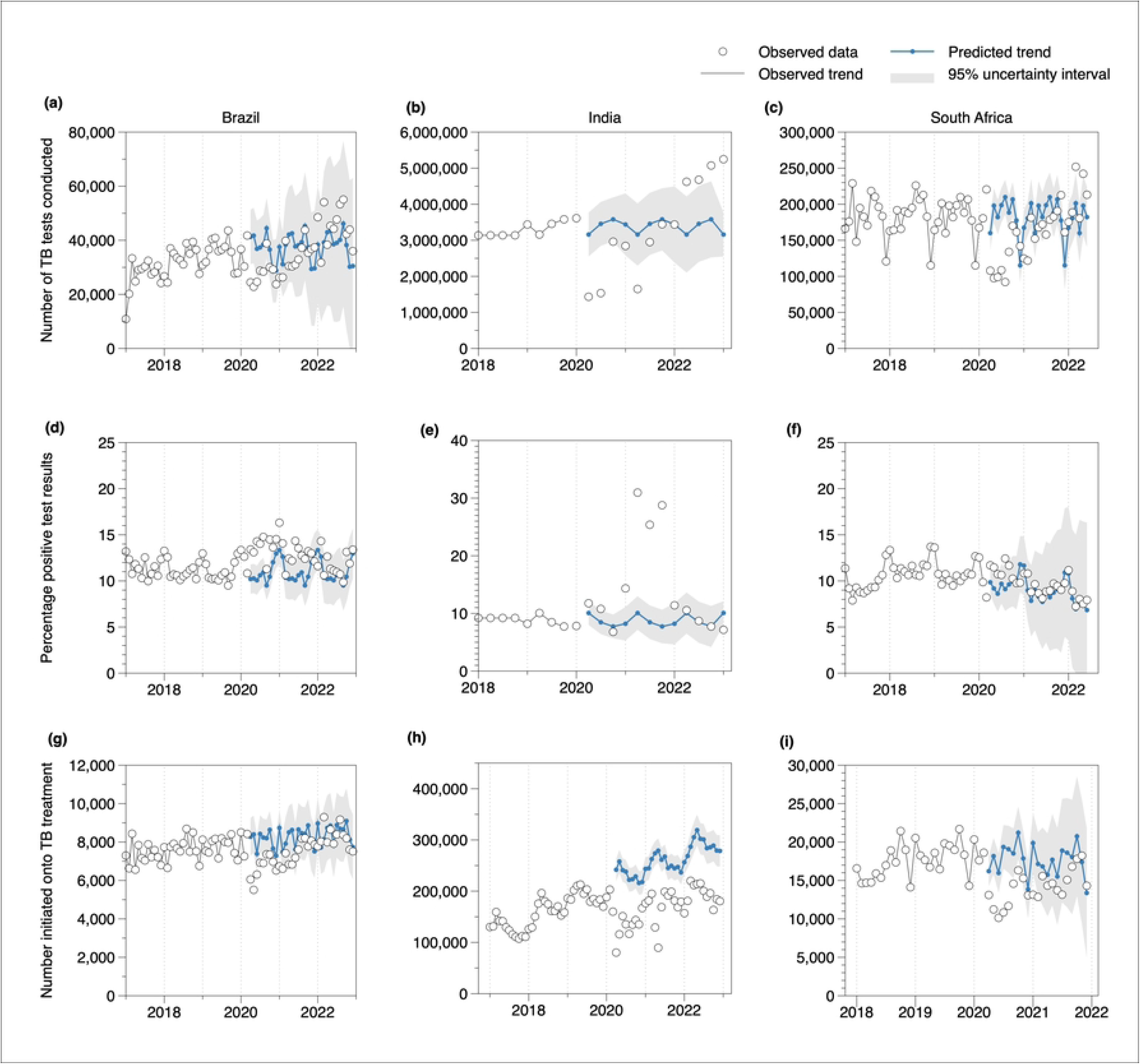
Time series data and ARIMA model forecast results for TB testing and treatment initiation in Brazil, India, and South Africa. White dots: observed TB test data; connected blue dots: ARIMA model forecasts; grey shaded areas: 95% prediction intervals; only annual TB test data were available for 2018 in India, therefore quarterly estimates were estimated from annual figures (assuming no seasonal variation).

**Figure 2.**
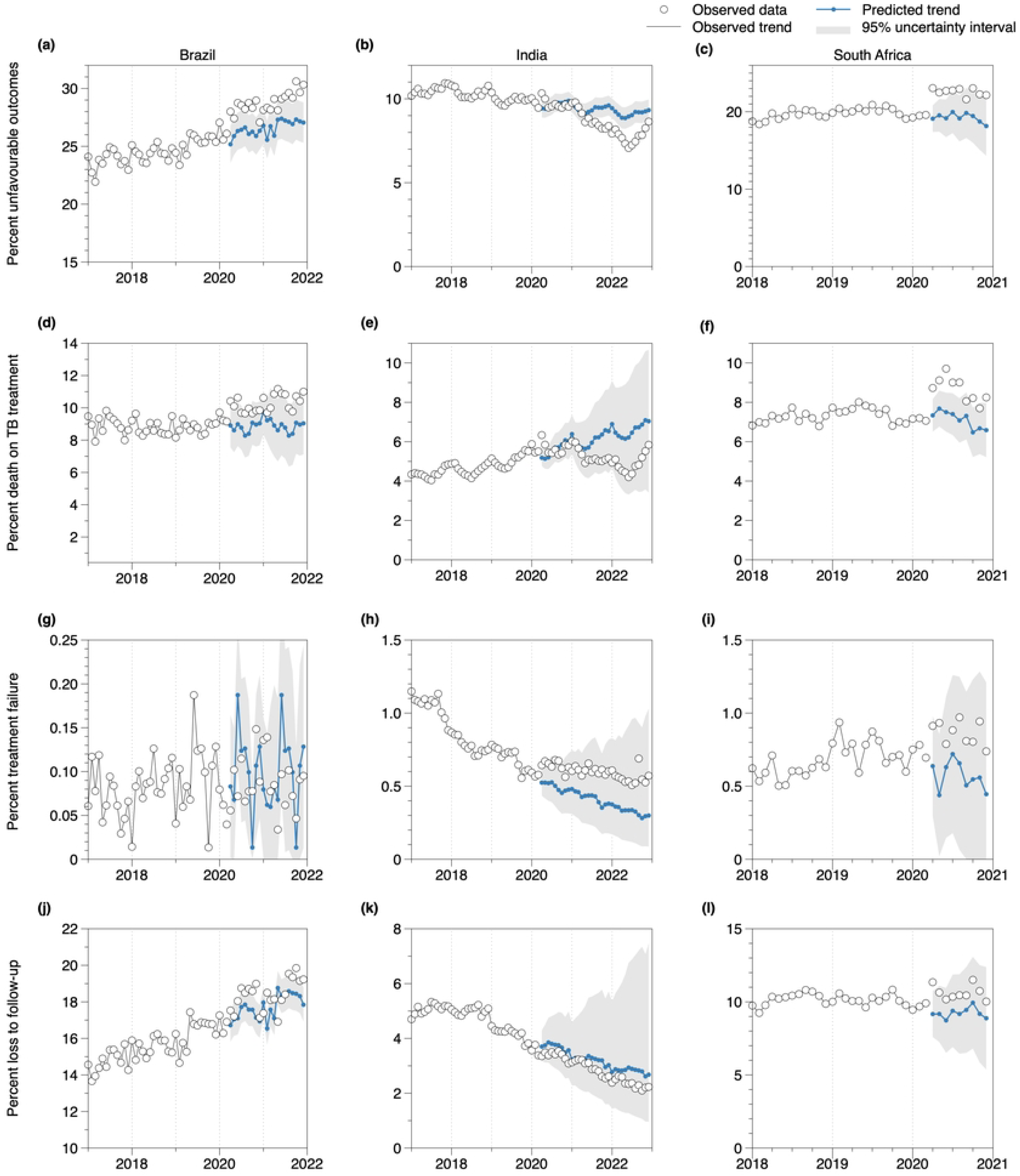
Time-series data and ARIMA model forecast results for unfavourable TB treatment outcomes in Brazil, India, and South Africa. White dots: observed TB test data; connected blue dots: ARIMA model forecasts; grey shaded areas: 95% prediction intervals

### TB testing, test positivity and treatment initiations during the COVID-19 pandemic

We found considerable reductions in the number of TB tests conducted during the first year of the COVID-19 pandemic in all three countries (Figure 1a-c). Between April and December 2020, the number of TB tests were lower by 24.3% (95% uncertainty interval: 8.4%; 36.6%) in Brazil, 27.8% (19.8%; 34.8%) in India and 32.0% (28.9%; 34.9%) in South Africa compared with predictions from the pre-COVID-19 period (Figure 3a). In 2021, reductions in the number of TB tests conducted were less severe compared with 2020 (Figure 1a-c). In 2022, all three countries—Brazil, India, and South Africa—surpassed the predicted number of bacteriological tests conducted during the pre-COVID period (Table S1). Notably, data from India suggested a substantial increase in the number of TB tests conducted relative to predicted values (Figure 1b).

**Figure 3.**
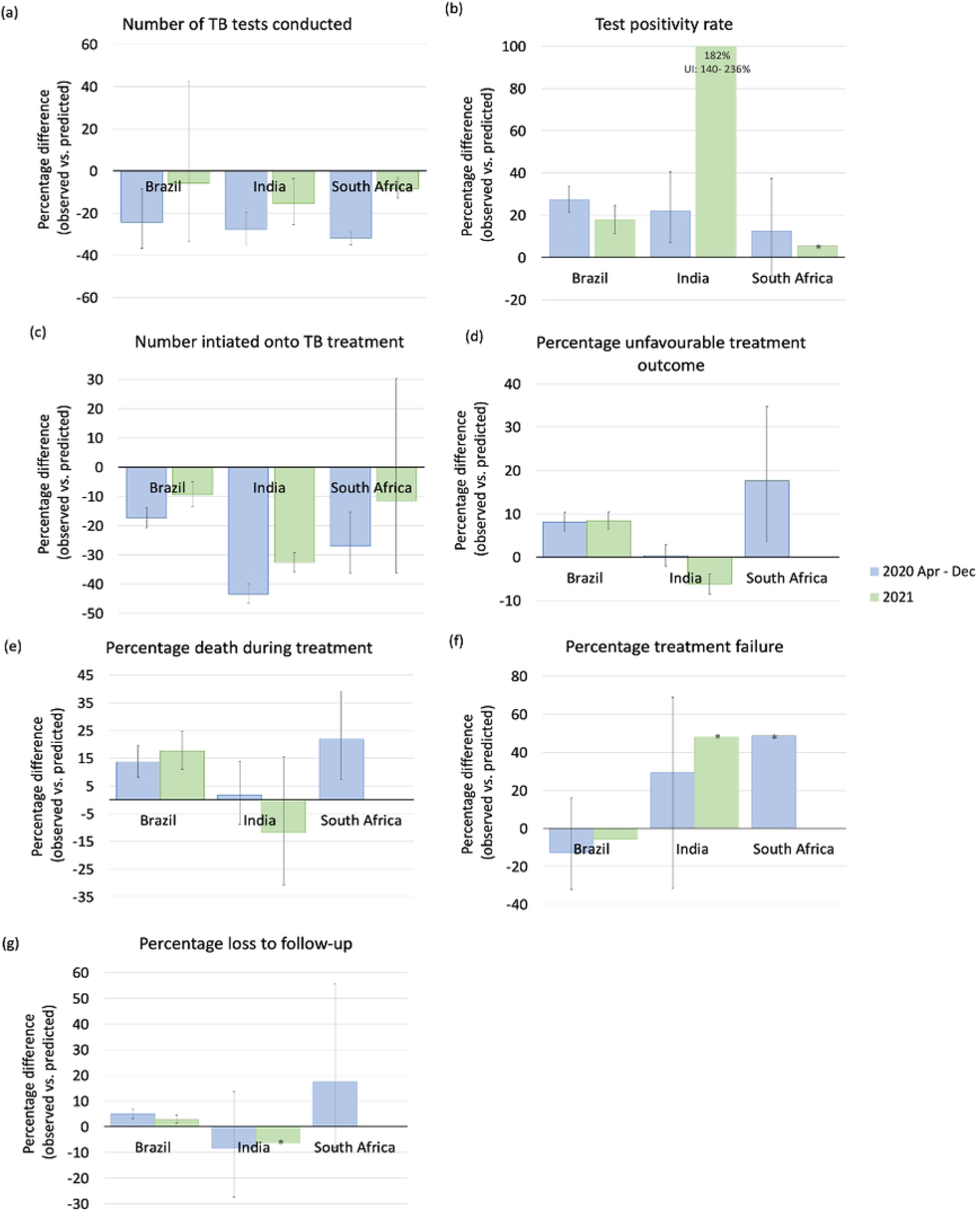
Average annual percentage differences between observed and predicted TB indicator values for the periods April-December 2020 and January-December 2021 in Brazil, India and South Africa. Error bars: 95% uncertainty intervals; negative percentage differences represent reductions in observed vs. predicted values; *Data bars without error bars show data for which uncertainty intervals could not be estimated as a log-transformation was used to prevent the ARIMA model from forecasting negative values.

Between April and December 2020, TB test positivity (Figure 1d-f) was 27.4% (21.4%; 33.8%) higher in Brazil, 22.0% (7.1%; 40.7%) higher in India and 12.5% (−8.6%; 37.4%) higher in South Africa compared with predictions from the pre-COVID period (Figure 3b). In India, a substantial increase in test positivity was observed in 2021, the year of the highest COVID-19 disease burden in the country. This was then followed by a decline to pre-COVID-19 levels in 2022 (Figure 1e).

The numbers of individuals initiating TB treatment (Figure 1g-i) were 17.4% (13.9%; 20.6%) lower than predicted in Brazil, 43.4% (39.8%; 46.4%) in India, and 27.0% (15.2%; 36.3%) in South Africa (Figure 3c). Reductions in treatment initiations were less severe in 2021 in all three countries. Additional analysis using data for South Africa showed that declines in TB treatment initiations were consistent with reductions in TB testing (Figure S1).

### TB treatment outcomes during the COVID-19 pandemic

In South Africa and Brazil, we observed a relative increase in the percentage of unfavourable TB treatment outcomes (Figure 2a,c), in particular death during TB treatment (Figure 2d,f), in the early phase of the COVID-19 pandemic. We estimated that in 2020, the percentage of unfavourable TB treatment outcomes was 8.1% (5.9%; 10.4%) higher than predicted in Brazil and 17.7% (3.5%; 34.8) higher in South Africa, compared with same-year predictions during the pre-COVID-19 period (Figure 3d). Annual percentages of death during TB treatment in 2020 were 13.7% (8.1%; 19.7%) higher in Brazil and 21.8% (7.4%; 39.2%) in South Africa (Figure 3e). In South Africa, we found a tendency toward higher percentages of loss to follow-up and treatment failure (Figure 2i,j). In 2021, relative increases remained stable in Brazil (Figure 3d,e; no data available for South Africa).

In India, percentages of unfavourable treatment outcomes and TB deaths during treatment in 2020 were consistent with pre-pandemic trends. The estimated percentage difference was 0.4% (−2.1%; 2.9%) for unfavourable treatment outcomes and 1,7% (−8.9%; 14.1%) for TB deaths during treatment relative to predictions from the pre-COVID-19 period in India (Figure 3d). Unfavourable treatment outcomes were lower than predicted in 2021 (Figure 3d).

## Discussion

In this study, we used time-series modelling to estimate the extent of disruptions to TB healthcare services during the COVID-19 pandemic in Brazil, India, and South Africa. Our findings suggest that TB healthcare services were seriously impacted in 2020, the first year of the pandemic, with at least partial reconstitution in the following years.

Our analysis revealed substantial reductions in the number of bacteriological tests for TB conducted during 2020 in all three countries compared with predictions from the pre-COVID period. Our findings are consistent with other reports from the early phase of the pandemic in Brazil [28] and South Africa [19] highlighting considerable reduction in TB tests performed. Our results also align with earlier reports of reduced TB service provision in these countries, indicating that laboratories redirected health and human resources to COVID-19, as well as reduced access to services due to restricted movement, loss of wages, and the fear of stigma [9]. We observed less severe reductions in TB testing in 2021, and relative increases in 2022, suggesting a partial recovery of TB diagnostic service and/or a catch-up in TB diagnoses were made.

Along with reduced TB testing, our analysis revealed substantial reductions in the number of people who initiated TB treatment during the COVID-19 pandemic in all three countries. Using data for South Africa, we found that the extent of relative reductions observed for treatment initiations compared well with TB tests conducted, suggesting that reduced TB testing during the COVID-19 pandemic was a main driver of reductions in the number of people initiating TB treatment.

Trends in TB treatment initiation based on data obtained and used for this study compare well with those of country-level TB case notifications reported by the WHO (Figure S2) [1]. One notable exception are case notifications in South Africa in the first half of 2020, which showed a more pronounced decline than that reported by the WHO. These differences may reflect later data updates and were communicated with the South African National TB Programme; a data review is currently under way.

We found higher TB test positivity rates than predicted from the pre-COVID period in Brazil and India, and a tendency toward higher test positivity in South Africa, consistent with a preliminary report by the South African National Institute for Communicable Diseases published in May 2020 [18] and a retrospective analysis of data from a hospital setting in India [29]. Higher test positivity rates than predicted could be due to differential healthcare seeking, more selective TB testing among people with presumptive TB, and/or differential reporting during the COVID-19 pandemic [30].

We found higher percentages of unfavorable treatment outcomes compared with predictions in the early phase of the COVID-19 pandemic, including for death during TB treatment. Increases in unfavourable treatment outcomes and a significant increase in death rates were previously reported for Eswatini (neighbouring South Africa) during the COVID-19 period [31] and an earlier time-series analyses found that COVID-19 caused a decline in TB cure rates in Brazil [16]. The rise in TB-related deaths among individuals undergoing TB treatment during the pandemic may be attributable to delays in TB diagnosis leading to more advanced and severe disease by the time treatment is initiated, as well as insufficient care during treatment. Higher numbers of deaths during TB treatment could also, at least in part, be attributed to COVID-19 [32].

Overall, the findings of this study are consistent between the three countries. However, we found notable differences. In contrast to Brazil and South Africa, model estimates for India do not suggest a notable recovery in treatment initiations in 2021, corresponding with the later peak of the COVID-19 crisis in the country. Furthermore, India witnessed more substantial increases in test positivity, reaching values of up to 30% in 2021 from 10% in prior years. Surprisingly, unlike for Brazil and South Africa, we did not estimate a relative increase in unfavorable treatment outcomes including deaths during TB treatment in India. Consistent with previously published reports [33], our data show a decline in TB deaths in India during the years 2020-2022 (Figure 2e), a reversal of trends observed in previous years. Whether challenges in reporting deaths and other adverse outcomes might serve to explain this, is currently not known. Alternatively, interventions implemented in India before and during the pandemic to support TB patients may be a reason why treatment outcomes in India did not worsen as much during the pandemic compared with other countries. Examples of these interventions include home delivery of TB medications, decentralized “drug refill facilities” in cities, involvement of commercial pharmacies in maintaining drug supplies, and the introduction of the ‘TB Aarogya Sathi’ app, a digital solution to enable direct interactions between TB patients and healthcare providers [34–36].

This study has several limitations. We evaluated numbers of bacteriological tests performed but were unable to investigate trends in the number of people accessing TB care. We are therefore unable to determine whether reductions in testing were attributable to reductions in care-seeking or shortages of TB tests. The use of cross-sectional aggregated data means that we could not link testing and treatment data on an individual level and could not investigate delays in healthcare service provision. We estimated differences in TB healthcare service indicators relative to pre-COVID trends but were unable to examine underlying factors and mechanisms that may have contributed to these differences. We are therefore unable to disentangle underlying causes. Most notably, we were unable to better understand and explain the considerable changes in tests conducted and test positivity in India. The accuracy, completeness, and timeliness of the data used in this study may be affected due to reduced staffing, increased workload during the pandemic affecting routine collection and reporting. Limited data points, statistical uncertainty, and random error in data from the pre-COVID period may have affected the accuracy of predicted (forecasted) trends in the indicators, affecting our ability to estimate relative differences during the pandemic period at varying degree. Finally, other external factors unrelated to the pandemic, for example increases in diagnostic coverage or in treatment initiations during the pre-pandemic period may have added uncertainty to model forecasts and thus have affected our findings.

In conclusion, our analysis reveals considerable disruptions to TB healthcare services in Brazil, India, and South Africa occurring during the early phase of the COVID-19 pandemic, including severe reductions in the number of TB tests performed and of TB treatment initiations during the early phase of the COVID-19 pandemic. We also estimated higher rates of unfavourable treatment outcomes, in particular TB deaths during treatment, in Brazil and South Africa. Our findings suggest at least partial recovery of services in the second and third year of the pandemic.

The substantial impact of the COVID-19 measures on TB healthcare service provision underscores the need for mitigation strategies to alleviate these detrimental effects. Especially in TB high-burden countries, plans for future pandemic preparedness should entail well-defined measures to prevent health-care service disruptions for TB and other diseases. This should include measures to preserve access to care during lockdowns as well as adequate staffing, and resources to sustain diagnostic and treatment services during pandemic times. As future pandemics are not unlikely, progress in ending the global TB pandemic in the forthcoming decades may depend on such efforts.

## Data Availability

Data on programmatic TB indicators used in this study were obtained from Departments of Health/National TB Programmes and country-level laboratory services, which are the custodians of the data. We are not permitted to publicly release these data on behalf of these institutions. We provide detailed information on data sources and availability in the manuscript. While enquiries to access the data must be made to the custodians, the authors of this study are willing, upon reasonable request to assist with contacting the respective institutions.

## Acknowledgments

We thank the Ministry of Health in Brazil, the Ministry of Health and Family Welfare in India, the TB Think Tank (Task Team: Epidemiology, Modelling and Health Economics) and the National Department of Health in South Africa as well as the National TB Programmes in all three countries for supporting this study. We are grateful to Nimalan Arinaminpathy (World Health Organization, Global TB Programme), Claudia Denkinger (Heidelberg University Hospital) and Cari van Schalkwyk (South African Centre for Epidemiological Modelling and Analysis) for helpful feedback to this manuscript.

## Supporting information

*Table S1. Average differences (%) between observed and predicted TB indicator values for the period April 2020 - December 2020, January 2021- December 2021 and January-December 2022 in Brazil, India, and South Africa. Negative values represent observed values lower than what was predicted*.

***Figure S1. Monthly average differences (%) between observed and predicted values for the number of TB tests conducted and TB treatment initiations in South Africa.*** *Negative values represent observed values lower than what was predicted*.

***Figure S2.*** ***Annual number of TB case notifications reported in Brazil, India and South Africa.** Blue lines represent WHO published data and the purple lines show the TB data we received for this study from the relevant TB programmes. WHO data was extracted from the 2022 Global Tuberculosis report*.

